# Optically Pumped Magnetoencephalography in Epilepsy

**DOI:** 10.1101/2019.12.10.19014423

**Authors:** Umesh Vivekananda, Stephanie Mellor, Tim M Tierney, Niall Holmes, Elena Boto, James Leggett, Gillian Roberts, Ryan Hill, Vladimir Litvak, Matthew J. Brookes, Richard Bowtell, Gareth R Barnes, Matthew C Walker

**Author notes:** Corresponding author: Dr Umesh Vivekananda; Department of Clinical and Experimental Epilepsy, Institute of Neurology, Queen Square, London WC1N 3BG, UK.

## Abstract

Our aim was to demonstrate the first use of Optically Pumped Magnetoencephalography (OP-MEG) in an epilepsy patient with unrestricted head movement. Current clinical MEG uses a traditional SQUID system for recording MEG signal, where sensors are cryogenically cooled and housed in a helmet in which the patient’s head is fixed. Here we use a different type of sensor (OPM), which operates at room temperature and can be placed directly on the patient’s scalp, permitting free head movement. We performed two 30 minute OP-MEG recording sessions in a patient with refractory focal epilepsy and compared these with clinical scalp EEG performed earlier. OP-MEG was able to identify analogous interictal activity to scalp EEG, and source localise this activity to an appropriate brain region. This is the first application of OP-MEG in human epilepsy. Future directions include simultaneous EEG/OP-MEG recording and prolonged OP-MEG telemetry.

## Introduction

Magnetoencephalography (MEG) is a non-invasive brain imaging technique that gives a unique window into whole brain function. In the field of epilepsy, MEG has been the source of much interest due to significant technical advantages it possesses over the ‘gold standard’ non-invasive technique for functional recording; scalp electroencephalography (EEG). Unlike EEG, the MEG signal is not affected by the smearing effects of skull and scalp and has better immunity to muscle artefact, both of which enable a greater ability to detect epileptic activity^1^. MEG has been shown to provide useful additional information to EEG, which significantly contributes to patient selection, focus localization and long-term seizure freedom after epilepsy surgery^2,3,4^. However use of MEG in epilepsy thus far has been limited to the pre-surgical evaluation of patients with drug-refractory epilepsies. This is because current clinical MEG uses an array of cryogenically-cooled sensors termed superconducting quantum interference devices or SQUIDs, placed around the head^5,6^, making MEG systems cumbersome and restrictive as sensor positions are fixed in a limited selection of helmet sizes. Consequently, any motion of the head relative to the sensors e.g. during a seizure, affects MEG signal quality in spite of movement compensation algorithms applied to it^7^. This means that MEG recording sessions are usually brief (1-2 hours) when compared to EEG telemetry that can last several days.

Here we describe the first use of an optically pumped magnetometer (OP) MEG system^8^ in a case of medically refractory focal epilepsy. OP-MEG utilises novel quantum sensors (optically-pumped magnetometers or OPMs) that do not rely on superconducting technology but on the transmission of laser light through a vapour of spin-polarised rubidium atoms. The use of OPMs in epilepsy has already been demonstrated in rodent models^9^. Crucially, OPM sensors can be worn directly on the head, allowing the subject to move within a magnetically shielded environment whilst being scanned. A recent OPM study showed sensory motor signals robust to subject movement of 20cm^8^. In addition, as the magnetically sensitive volume within the OPM sits just 6mm from the scalp surface by comparison to roughly 4cm in cryogenic MEG, the magnetic field strengths measured due to cortical sources are typically 4 times greater in adults^10,11^, since magnetic field strength decreases with distance. The intrinsic noise level of the most recent OPMs is comparable to that SQUIDs (∼10ft/sqrt(Hz))^8^.

In this study we demonstrate, for the first time in humans, that interictal epileptiform activity can be recorded using OPMs. The localization of this activity agrees with that of other modalities. OP-MEG therefore promises a combination of high spatiotemporal accuracy during brain recording (with minimal effects of motion or muscle artefact) and practicality, with subjects able to move naturally during recordings.

### Case

Our test case was a 47-year-old woman who had Haemophilus meningitis associated with status epilepticus aged 18 months. After a period of seizure freedom, she developed focal seizures with loss of awareness, refractory to a number of antiepileptic drugs, and she is currently experiencing up to 10 seizures a day. Repeated diagnostic EEG demonstrated epileptic spikes, polyspikes, and spike and wave patterns indicative of a right posterior quadrant focus for the epilepsy. MRI demonstrated widespread parieto-occipital damage of cortex and white matter, indicative of a previous ischaemic insult.

## Method

Prior ethical approval for this study was granted by The Medicines and Healthcare products Regulatory Agency. Informed consent was obtained from the patient to participate in the study. Band-pass filtering between 1 and 70Hz was applied to the EEG and OP-MEG data. Prolonged EEG monitoring (5 days) was performed with a MedTronic system with a 21-channel setup using the international 10-20 electrode system and average reference montage. The patient subsequently underwent two separate recording sessions using OP-MEG, each lasting 30 minutes. For both recording sessions, we used remote reference sensors placed near the patient to record, and eventually regress out, environmental magnetic noise. The first session involved the patient lying on a bed with her head resting on a bespoke plinth, which housed 8 first-generation OPMs (Figure 1A2, left) for recording brain activity and 6 sensors for reference measurement. The second session was performed with the subject wearing a bespoke 3D-printed scanner-cast (Figure 1A1) to hold 15 second-generation OPMs (Figure 1A2, right). The scanner-cast design was informed by a previously acquired anatomical 3 T MRI scan^12^. These casts provide a rigid sensor mount with respect to the individual anatomy and ensures that any movement artefacts are common across sensors. The positions of the sensors in relation to the brain could be ascertained directly from the digital scanner cast design (Figure 1B). Source reconstruction of an average of interictal spikes (manually identified by an experienced neurophysiologist) was performed using dipole fitting within Fieldtrip software (http://fieldtriptoolbox.org) (Figure 1D-F). We used the rising phase to maximal peak of spike for reconstruction. Both recordings were performed within a magnetically shielded room and with the subject sitting between two biplanar coils separated by 1.5 metres, which were used to remove remnant static magnetic fields^13^. This allowed the patient to move her head naturally whilst reclining in a chair.

**Figure 1.**
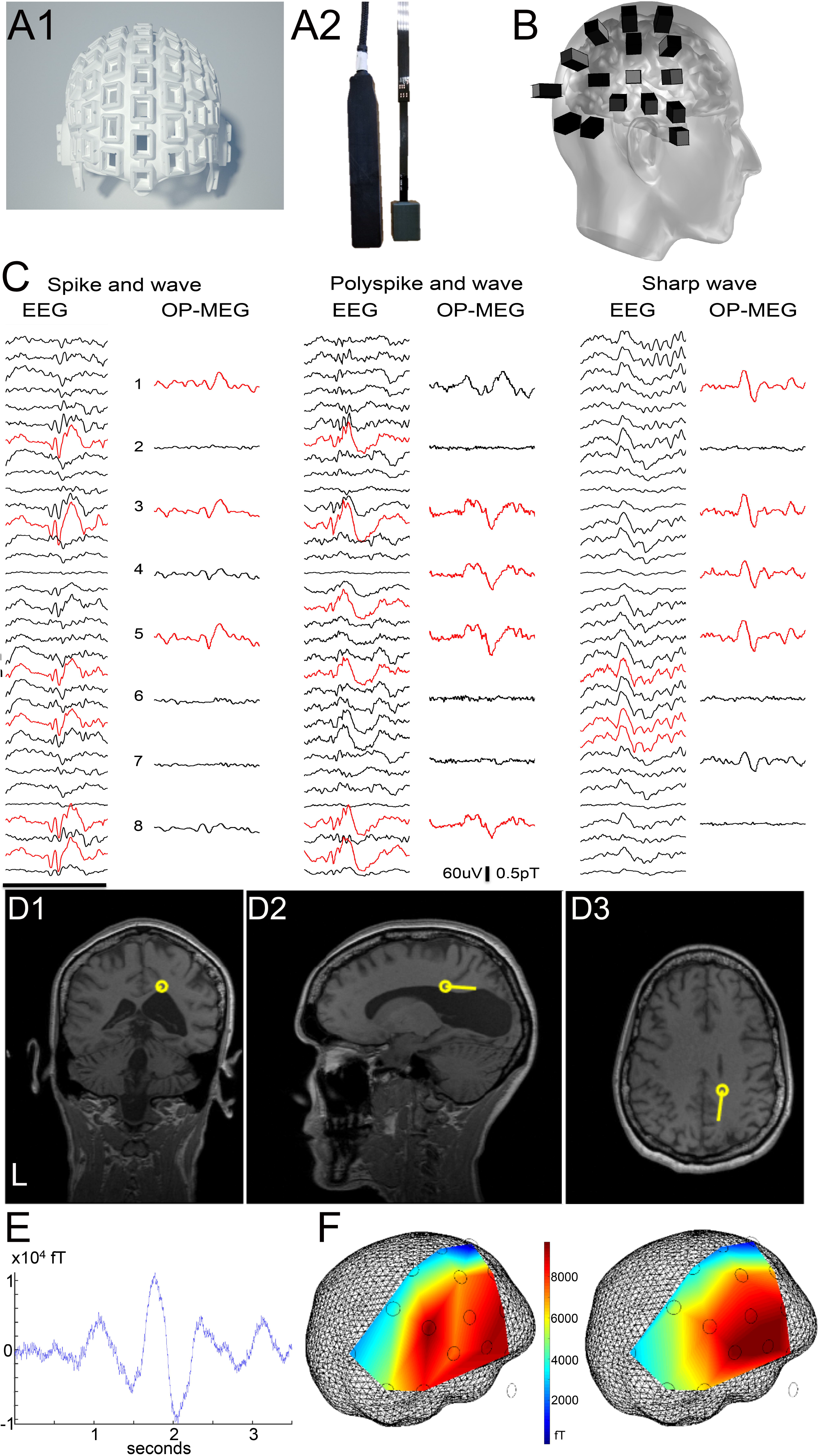
Use of OP-MEG in epilepsy. A1. Rendered image of 3D scanner-cast A2. First generation (left) and second generation (right) OPM sensor B. Position of OPM sensors on the scalp C. Example traces of typical epileptiform activity from scalp EEG and OP-MEG (first recording session using Gen1 sensors), including spike and wave, polyspike and wave, sharp wave. Red denotes channels with highest amplitude spikes. EEG labelled with average reference montage; Black bar = 1 second D. Source reconstruction of average interictal spike activity using OP-MEG; D1 coronal, D2 sagittal and D3 axial using dipole fit. E. Example average OP-MEG sharp wave used for reconstruction F. Field map (measured left and modelled right) corresponding to the peak of the epileptic spike with sensor positions shown in relation to the inner skull mesh,

## Results

During EEG telemetry the patient exhibited frequent interictal activity including sharp waves, spikes and polyspikes, on average 30/hour (examples in Figure 1C). These were maximal in channels T8 and Right Sphenoidal. She also had four seizures heralded by right fronto-centro-temporal spikes and subsequent generalised attenuation of EEG activity. During the two OP-MEG recording sessions, we were able to identify the same interictal patterns, on average 21 spikes/30 minutes. The patient suffered no seizures during OP-MEG recording. Using 19 spikes recorded in one session, we localised the epileptiform activity to the previously EEG identified abnormal right posterior quadrant, assumed to be the epileptogenic focus (Figure 1D). She since underwent resective surgery of this region with a reduction in her seizure frequency.

## Conclusion

We have demonstrated the first use of OP-MEG in an epilepsy patient with unrestricted head movement. OP-MEG detected forms of abnormal interictal activity seen on clinical EEG (spikes, polyspikes, spike and wave) demonstrating similar morphology, and produced consistent localisation of spike activity. One limitation of this study has been that EEG and OP-MEG were not acquired at the same time. However, this study is an important proof of concept step in the use of OP-MEG for epilepsy. Another limitation is that the OP-MEG sensors were concentrated around the region of interest when recording; i.e. right posterior quadrant. Although this introduces some bias as compared to whole head coverage, an inherent advantage of MEG over EEG is overcoming this inverse problem of source localisation. Further development of the technique will include simultaneous 30 channel OP-MEG and EEG recording, which has already been demonstrated in healthy subjects^14^ and prolonged EEG/OP-MEG recording over several hours with video monitoring. Work is also underway in developing OP-MEG for children with refractory epilepsy where motion tolerance is of even more importance. In addition, a new large OP-MEG recording suite (3 x 4 metres) is being completed at Wellcome Centre for Human Neuroimaging, UCL, with improved low frequency shielding to allow a larger area for the patient to move in whilst recording. OP-MEG has the potential to improve the quality of functional imaging within epilepsy diagnostics and finally translate MEG into a readily clinically available tool.

## Data Availability

NHS Ethics have requested that this patient data would not be available for sharing

## Acknowledgments

We would like to acknowledge the patient that participated in this is study. This work was supported by the Department of Health’s National Institute for Health Research University College London/University College London Biomedical Research Centre and Epilepsy Research UK.

This work was funded by a Wellcome collaborative award to Barnes, Brookes and Bowtell, which involves a collaborative agreement with the OPM manufacturer QuSpin. SM is supported by the EPSRC-funded UCL Centre for Doctoral Training in Medical Imaging (EP/L016478/1). The Wellcome Centre for Human Neuroimaging is supported by core funding from the Wellcome (203147/Z/16/Z). There are no further conflicts of interest to disclose. We confirm that we have read the Journal’s position on issues involved in ethical publication and affirm that this report is consistent with those guidelines.

